# Systematic review and meta-analysis of the prevalence of substance use among adolescents in South Africa

**DOI:** 10.1101/2025.09.18.25336046

**Authors:** Carrie Brooke-Sumner, Lauro Estivalete Marchionatti, Zeina Mneimneh, Nadine Harker, Catherine O. Egbe, Dan Jenkins, Noluthando Mpisane, Meghan Mosalisa, Yeukai Chideya, Nuhaa Holland, Giovanni Salum, Jason Bantjes

## Abstract

**Background:** Substance use among young people is a growing public health concern in South Africa (SA), with implications for short- and long-term mental health, and education and development. A lack of use data is amongst several challenges impeding progress to support young people. To address this gap, this study aimed to synthesize published prevalence data on substance use among adolescents under 19 years of age in SA.

**Methodology:** We conducted a systematic review and meta-analysis following PRISMA guidelines. Searches were run across PubMed, PsycINFO, Web of Science and Scielo. Studies reporting original SA prevalence data in community- or school-based samples were included. The Joanna Briggs Institute checklist for prevalence studies was used to assess study quality. Meta-analyses were performed in R, using random effects models, and heterogeneity was assessed using I^2^ statistics and meta-regression.

**Results:** Thirty studies met inclusion criteria, representing 202 prevalence estimates (n=120 041, mean age 12.09-19 years old) across the nine provinces of the country. The most commonly reported substances used were alcohol (37%, 95%CI=30.36-48.39), tobacco (25.66% 95%CI=17.12-34.19), and cannabis (12.63% 95%CI=7.38-17.88) with lifetime prevalence of any substance use of 17.11% 95%CI=13.51-20.7239. Similarly, 12-month data indicated high exposure levels: alcohol 33.17% (95%CI=19.51-46.82), tobacco 15.82% (95%CI=8.73-22.90), and cannabis 8.27% 95%CI=3.96-12.57. However, substantial heterogeneity across studies was detected.

**Conclusion:** High exposure to substances in South Africa, especially alcohol, tobacco, and cannabis, underscores the urgent need for nationally representative surveillance and evidence-based prevention efforts tailored to adolescents, particularly in early intervention to reduce progression of substance use to disorder.

## Introduction

Young people are the foundation of a developing society [1], constituting more than a third of the population in South Africa [2] Protecting the health and wellbeing of young people is crucial for these countries to achieve health and development goals. Equally importantly, childhood and adolescence are critical developmental stages, in terms of cementing personal identity and integrating into social environments [3]. Maturation is required in this time for healthy decision-making, impulse control, and emotional regulation and there is potential for increased risk-taking behaviour during this phase [4, 5]. It follows that young people are particularly vulnerable to stresses (e.g. peer pressure, and environmental influences [6] social and cultural norms) with exposure to substance use and adverse experiences having a lasting impact on their developmental trajectory [1]. Consequences of this vulnerability are borne out, as early initiation of substance use is associated with higher risk of developing substance use disorders, mental health conditions, and chronic diseases later in life [7]. This in addition to impaired cognitive development [8], risk for lower academic achievement [9], and increased likelihood of engaging in risky behaviors e.g. unprotected sex [5, 10] which further exacerbate the overall burden of substance use on young people’s health and development. Substance use in young people is a global and African public health concern [11] with substantial public health burden, which is contributing increasingly to Years Lived with Disability (YLDs) [12, 13]. About a quarter of young people have used substances in their lifetime and this comes with physical and socioeconomic costs [14]. Tobacco and alcohol remain the most commonly used substances, and prevalence and patterns of use vary widely across regions, influenced by cultural norms, factors that affect access to substances, socioeconomic factors, and policy environments [15, 16].

Substance use in young people in South Africa has been a growing public health concern, driven by these socioeconomic cultural and environmental factors and stressors experienced by young people [17–19], including exposure to violence [20]. For more than a decade there have been calls to address harms of substance use among South African youth [11] notably binge drinking, tobacco and cannabis use and the harms of newer synthetic drugs and over-the-counter medications. The South African National Youth Risk Behaviour Survey waves 2002, 2008, 2011 and the Global Youth Tobacco Survey waves 1998, 2001, 2003 and 2011 have underscored the high prevalence of alcohol and tobacco use and increases in cannabis use over time, but the country is lacking robust national and provincial estimates for use in young people. Indicators of the burden of the trajectory of substance use to disorder for young people are indicated by the South African Community Epidemiology Network on Drug Use (SACENDU) routine surveillance data on national treatment admission trends [21]. Although the country has a National Drug Master Plan [22], challenges persist in scaling up prevention and intervention programmes, hinged on lack of evidence on patterns of use across provinces and nationally, with consequent lack of appropriate resource allocation and action.

With this background and to provide impetus and direction for evidence-based prevention for substance use and disorder, this study aimed to synthesize available data of the prevalence of substance use exposure among adolescents young people in South Africa. This review is part of the Child and Adolescent Mental Health Initiative in South Africa (CAMHISA) – a systems strengthening program that aims to strengthen the capabilities of mental health care providers, expanding access to high-quality care, and improving mental health outcomes for young people in South Africa. It stands alongside a review of prevalence of mental health conditions, which shows the landscape of child and adolescent mental health conditions in South Africa as a platform for implementation of treatment and prevention (Ref). The current review synthesizes data on exposure to substances (not substance use disorder, which is covered in the mental disorders prevalence review) to provide evidence for the extent of exposure to substances in South African youth. The methods used in this review draw on related reviews conducted in Greece and Brazil (Koumoula et al., 2023; Marchionatti et al., 2024).

## Methods

The review is based on a nationwide South African review on child and adolescent mental health, conducting a meta-analysis of studies specifically reporting substance use. For the full nationwide review, refer to preprint. Methods are based on previous national efforts in Brazil and Greece, with data extraction and appraisal followed guidance of a widely referred meta-analysis on prevalence of child and adolescent mental disorders [23]. The protocol was registered on the Prospero Database (CRD42024619173). Findings are presented here in line with the Preferred Reporting Items for Systematic Reviews and Meta-Analysis (PRISMA) statement [24].

### Search Strategy

The overall search strategy for the landscape review of child and adolescent mental health in South Africa used English search terms covering children and adolescents, South Africa, mental health, mental disorders, suicide and substance use (see supplementary materials Table S1). Searches were developed and run in March 2024 and then updated in February 2025 for PubMed, PsycINFO, Web of Science, and Scielo.org (South Africa Collection) without restriction to language or date. To strengthen this approach and check for articles that could be missed in the other databases, we also performed a supplementary search on Google Scholar using general search phrases (for example, “substance use” AND (“child” OR “adolescent”) AND “South Africa”). We reviewed these results until reaching 50 consecutive Google scholar records (2 reviewers) without finding any that had not been previously identified by other databases. The reference lists of systematic reviews were also targeted for backward citation searching to find further potentially relevant studies.

### Data management

Search results were initially viewed in EPPI-Reviewer 4.0. This was used for deduplication, with the calibration index for detection set at 0.85, which has been demonstrated to provide a very low rate of false positives [25]. Subsequently, identified studies were uploaded to Rayyan, which was used for further deduplication with manual verification of possible duplicates, and data management.

### Screening procedures

To identify possible studies for inclusion, two researchers independently examined every record in Rayyan by title and abstract, applying the inclusion criteria below. Discussions and communication with a third reviewer were used to settle inclusion disagreements that arose. Secondary screening based on full-texts of studies was made by a single researcher, with discussion with the broader team if required. Studies needed to fulfil all of the inclusion criteria below to be included.

### Inclusion criteria

The criteria of the primary review: prevalence estimates reported for samples adolescents with average age under 19 in SA, gathered from community- and school-based surveys either from self- or proxy-report, published in which types of publications. The criteria for the specific review: the subset of studies on prevalence of substance use

1. Provide original SA data gathered from surveys with community- and/or school-based samples or epidemiological registers.
2. Report on a sample of adolescents under the age of 19, or if young adults were included in the study, a sample with a mean age under the age of 19 was included.
3. Include data gathered from informants (parents, guardians, teachers, etc.) and/or self-report data (children/adolescents).
4. Contain data on prevalence estimates for substance use.
5. In case of duplicate datasets, included data should be the earliest and/or most comprehensive report of the data in question
6. Have findings published in English in any of the following formats: journal articles, editorial letters if original data was reported, book chapters if original data was reported, or reports of national surveys.

Studies were excluded if they met any of the following criteria:

1. Only presented data from clinical or non-representative populations and those with characteristics unrepresentative of the broader community (e.g., adolescents living with HIV).
2. Included substance use data presented as absolute numbers rather than rates (i.e. no denominator).
3. Multi-country studies including South African data that could not be disaggregated and general population studies without data specifically for children and adolescents.

### Data extraction

Prior to full text screening and data extraction, a pilot test of screening and data extraction was conducted by each member of the research team applying the inclusion/exclusion criteria to a random selection of 10 publications. The research team then met to discuss agreement and adjustments to the data extraction tool were made accordingly. A RedCap-programmed data extraction form was used to extract data. This form enabled extraction of comprehensive information on study design, setting, procedures, and outcomes. Data was extracted by one reviewer for each source article. Data were exported from RedCap to Excel and checked for accuracy and completeness by a second reviewer who referred to the original publications. See Supplementary Table S1 for all extracted information from included studies.

### Quality assessment

Studies included in this review were assessed for methodological quality using the Joanna Briggs Institute (JBI) Critical Appraisal Checklist for Prevalence Studies [26]. This checklist evaluates key aspects of prevalence studies including sample representativeness, recruitment methods, data collection procedures, and statistical analyses. Quality assessment was not used as a basis for excluding lower quality studies which would not have aligned with the aims of this landscape review, but rather to enable us to provide an overall assessment of the quality of the body of evidence.

### Data synthesis and analysis

Meta-analysis was conducted for substances with 5 or more prevalence estimates (lifetime, 12-month). This number of estimates was required to enable meaningful synthesis and statistical reliability [27, 28]. Pooled prevalence rates calculations and meta-analysis were conducted using the metafor package in R [29].

## Results

The initial searches (17 219 records) identified 11 483 abstracts for review by title and abstract after removing duplicate records. An additional 27 studies were identified from scanning the refences in 30 systematic reviews identified giving a total of 11 510 titles and abstracts reviewed. Of these, 291 were selected for full-text review and 30 were included after applying all inclusion and exclusion criteria (see PRISMA flow chart in Figure 1 below). Three of the included studies reported secondary data from national youth risk surveys [30–32] so they were removed and the three original reports with the primary survey data were included in their place. The final number of included publications with substance use data was 30 (Table 1).

**Table 1.**
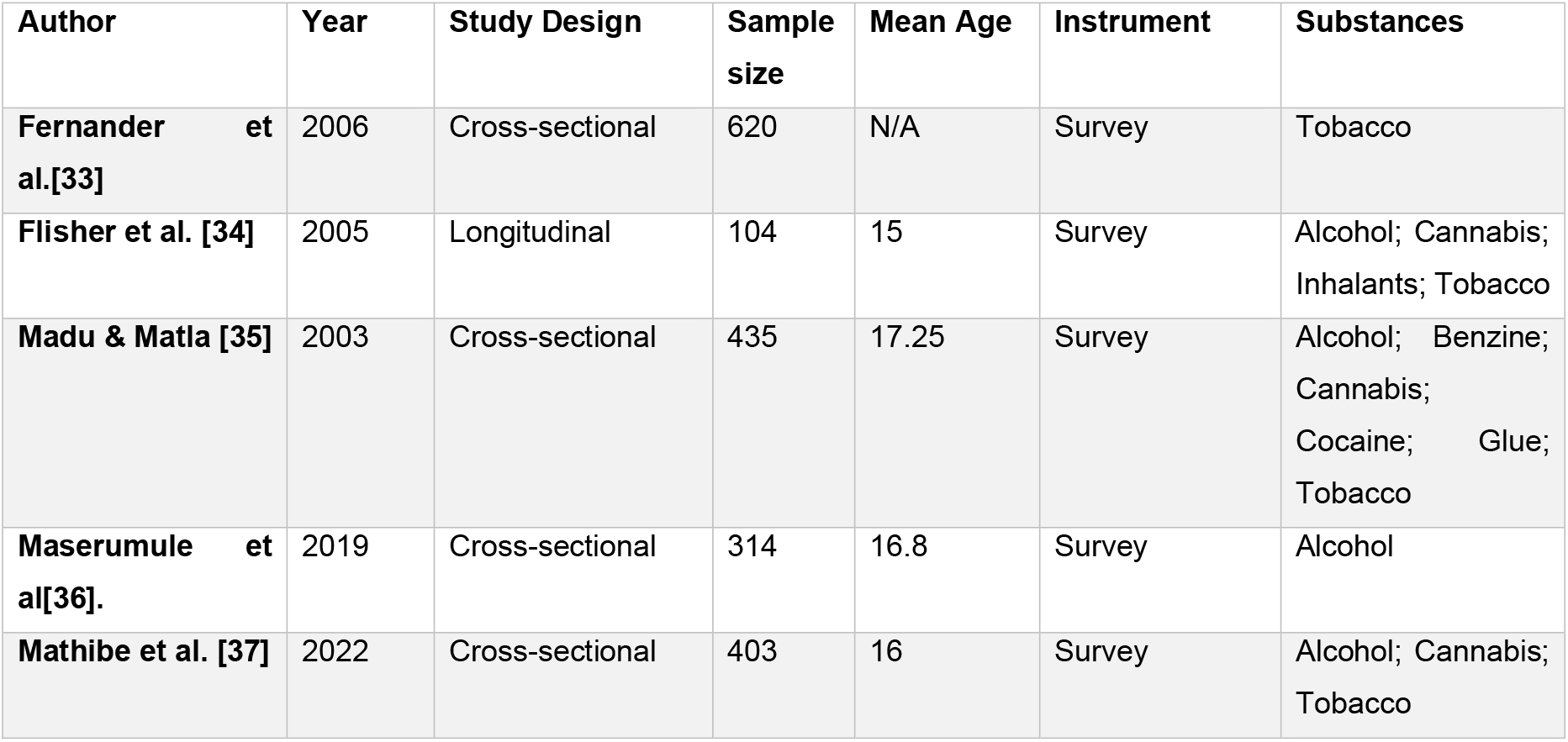
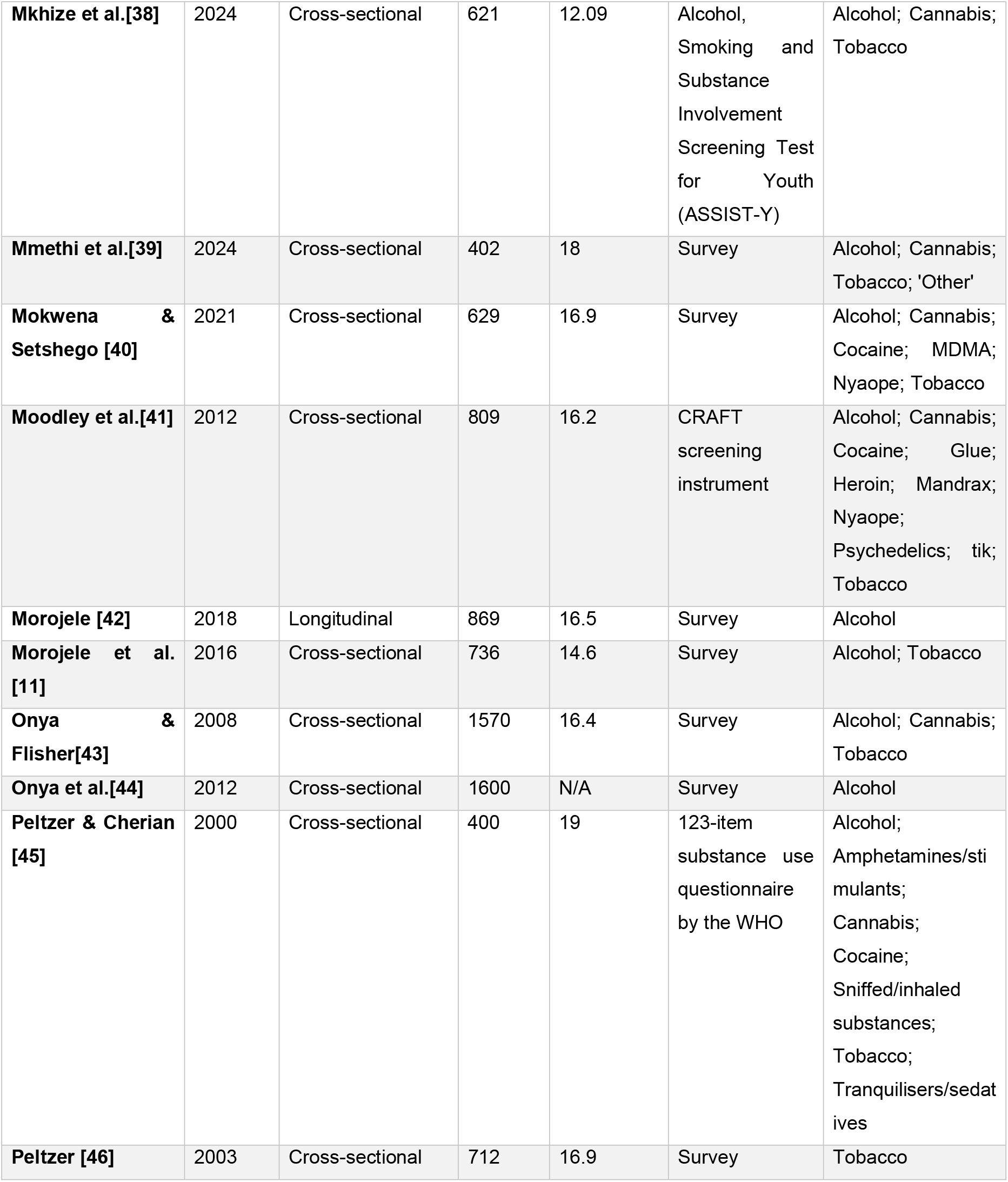
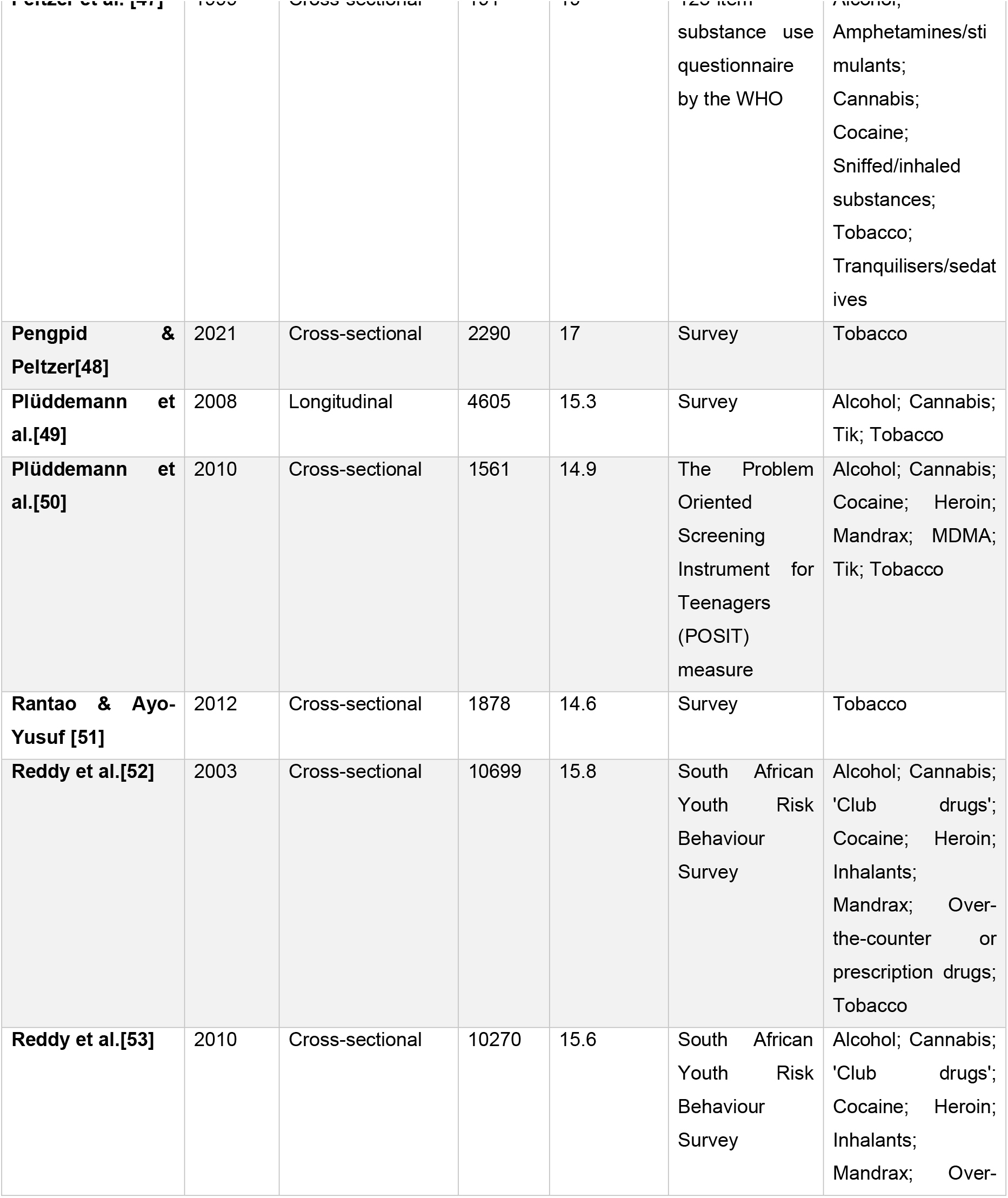
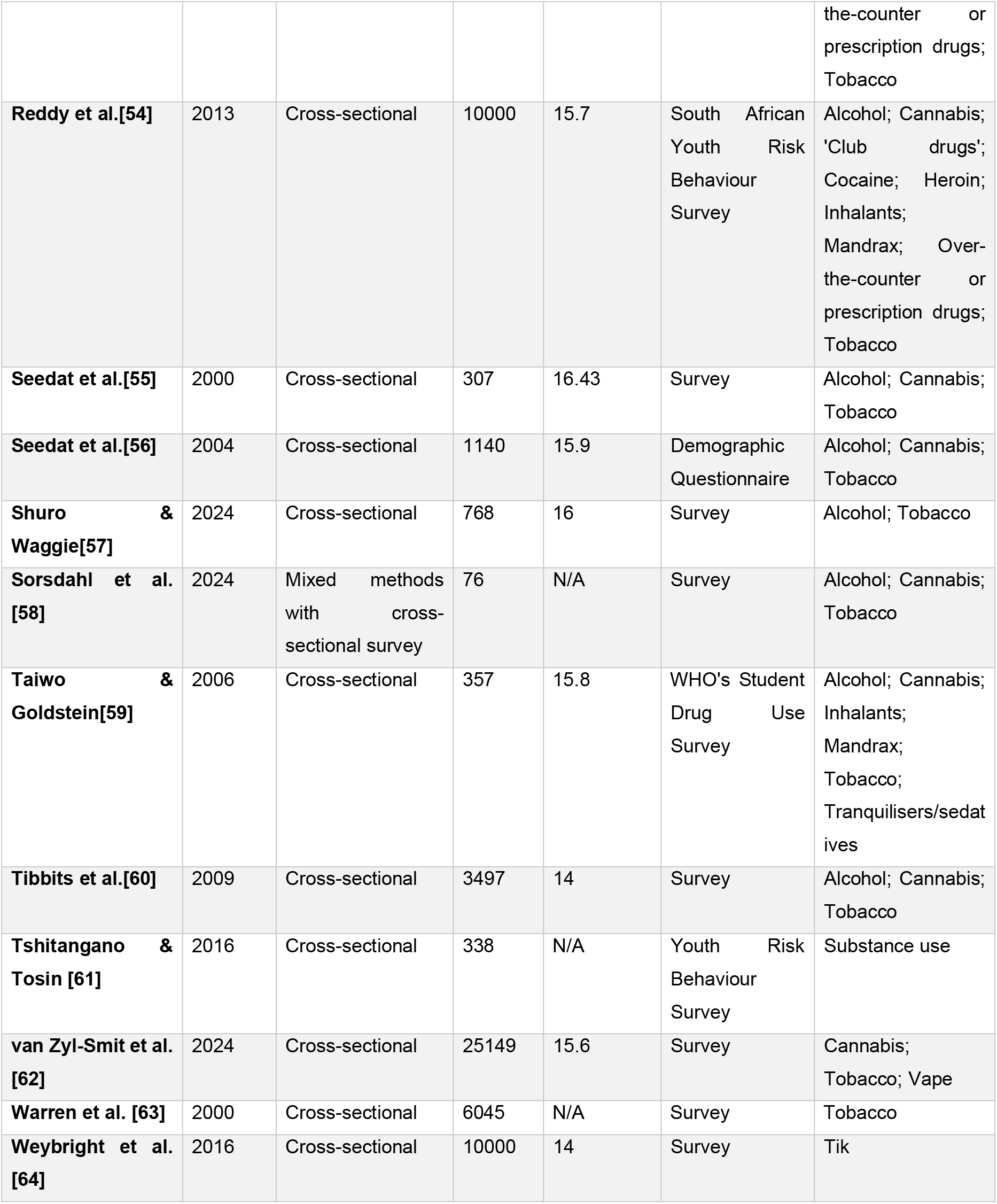
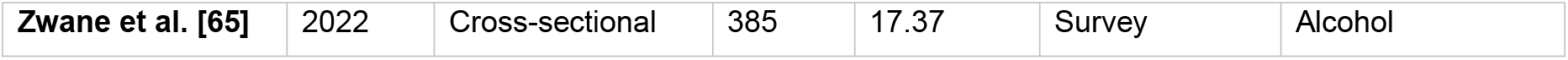
Summary of included studies.

| Author | Year | Study Design | Sample size | Mean Age | Instrument | Substances |
| --- | --- | --- | --- | --- | --- | --- |
| **Fernander et al.[33]** | 2006 | Cross-sectional | 620 | N/A | Survey | Tobacco |
| **Flisher et al. [34]** | 2005 | Longitudinal | 104 | 15 | Survey | Alcohol; Cannabis; Inhalants; Tobacco |
| **Madu & Matla [35]** | 2003 | Cross-sectional | 435 | 17.25 | Survey | Alcohol; Benzine; Cannabis; Cocaine; Glue; Tobacco |
| **Maserumule et al[36].** | 2019 | Cross-sectional | 314 | 16.8 | Survey | Alcohol |
| **Mathibe et al. [37]** | 2022 | Cross-sectional | 403 | 16 | Survey | Alcohol; Cannabis; Tobacco |
| **Mkhize et al.[38]** | 2024 | Cross-sectional | 621 | 12.09 | Alcohol, Smoking and Substance Involvement Screening Test for Youth (ASSIST-Y) | Alcohol; Cannabis; Tobacco |
| **Mmethi et al.[39]** | 2024 | Cross-sectional | 402 | 18 | Survey | Alcohol; Cannabis; Tobacco; 'Other' |
| **Mokwena & Setshego [40]** | 2021 | Cross-sectional | 629 | 16.9 | Survey | Alcohol; Cannabis; Cocaine; MDMA; Nyaope; Tobacco |
| **Moodley et al.[41]** | 2012 | Cross-sectional | 809 | 16.2 | CRAFT screening instrument | Alcohol; Cannabis; Cocaine; Glue; Heroin; Mandrax; Nyaope; Psychedelics; tik; Tobacco |
| **Morojele [42]** | 2018 | Longitudinal | 869 | 16.5 | Survey | Alcohol |
| **Morojele et al. [11]** | 2016 | Cross-sectional | 736 | 14.6 | Survey | Alcohol; Tobacco |
| **Onya & Flisher[43]** | 2008 | Cross-sectional | 1570 | 16.4 | Survey | Alcohol; Cannabis; Tobacco |
| **Onya et al.[44]** | 2012 | Cross-sectional | 1600 | N/A | Survey | Alcohol |
| **Peltzer & Cherian [45]** | 2000 | Cross-sectional | 400 | 19 | 123-item substance use questionnaire by the WHO | Alcohol; Amphetamines/stimulants; Cannabis; Cocaine; Sniffed/inhaled substances; Tobacco; Tranquilisers/sedatives |
| **Peltzer [46]** | 2003 | Cross-sectional | 712 | 16.9 | Survey | Tobacco |
| **Peltzer et al. [47]** | 1999 | Cross-sectional | 191 | 19 | 123-item substance use questionnaire by the WHO | Alcohol; Amphetamines/stimulants; Cannabis; Cocaine; Sniffed/inhaled substances; Tobacco; Tranquilisers/sedatives |
| **Pengpid & Peltzer[48]** | 2021 | Cross-sectional | 2290 | 17 | Survey | Tobacco |
| **Plüddemann et al.[49]** | 2008 | Longitudinal | 4605 | 15.3 | Survey | Alcohol; Cannabis; Tik; Tobacco |
| **Plüddemann et al.[50]** | 2010 | Cross-sectional | 1561 | 14.9 | The Problem Oriented Screening Instrument for Teenagers (POSIT) measure | Alcohol; Cannabis; Cocaine; Heroin; Mandrax; MDMA; Tik; Tobacco |
| **Rantao & Ayo-Yusuf [51]** | 2012 | Cross-sectional | 1878 | 14.6 | Survey | Tobacco |
| **Reddy et al.[52]** | 2003 | Cross-sectional | 10699 | 15.8 | South African Youth Risk Behaviour Survey | Alcohol; Cannabis; 'Club drugs'; Cocaine; Heroin; Inhalants; Mandrax; Over-the-counter or prescription drugs; Tobacco |
| **Reddy et al.[53]** | 2010 | Cross-sectional | 10270 | 15.6 | South African Youth Risk Behaviour Survey | Alcohol; Cannabis; 'Club drugs'; Cocaine; Heroin; Inhalants; Mandrax; Over- the-counter or prescription drugs; Tobacco |
| **Reddy et al.[54]** | 2013 | Cross-sectional | 10000 | 15.7 | South African Youth Risk Behaviour Survey | Alcohol; Cannabis; 'Club drugs'; Cocaine; Heroin; Inhalants; Mandrax; Over-the-counter or prescription drugs; Tobacco |
| **Seedat et al.[55]** | 2000 | Cross-sectional | 307 | 16.43 | Survey | Alcohol; Cannabis; Tobacco |
| **Seedat et al.[56]** | 2004 | Cross-sectional | 1140 | 15.9 | Demographic Questionnaire | Alcohol; Cannabis; Tobacco |
| **Shuro & Waggie[57]** | 2024 | Cross-sectional | 768 | 16 | Survey | Alcohol; Tobacco |
| **Sorsdahl et al. [58]** | 2024 | Mixed methods with cross-sectional survey | 76 | N/A | Survey | Alcohol; Cannabis; Tobacco |
| **Taiwo & Goldstein[59]** | 2006 | Cross-sectional | 357 | 15.8 | WHO's Student Drug Use Survey | Alcohol; Cannabis; Inhalants; Mandrax; Tobacco; Tranquilisers/sedatives |
| **Tibbits et al.[60]** | 2009 | Cross-sectional | 3497 | 14 | Survey | Alcohol; Cannabis; Tobacco |
| **Tshitangano & Tosin [61]** | 2016 | Cross-sectional | 338 | N/A | Youth Risk Behaviour Survey | Substance use |
| **van Zyl-Smit et al. [62]** | 2024 | Cross-sectional | 25149 | 15.6 | Survey | Cannabis; Tobacco; Vape |
| **Warren et al. [63]** | 2000 | Cross-sectional | 6045 | N/A | Survey | Tobacco |
| **Weybright et al. [64]** | 2016 | Cross-sectional | 10000 | 14 | Survey | Tik |
| **Zwane et al. [65]** | 2022 | Cross-sectional | 385 | 17.37 | Survey | Alcohol |

**Figure 1:**
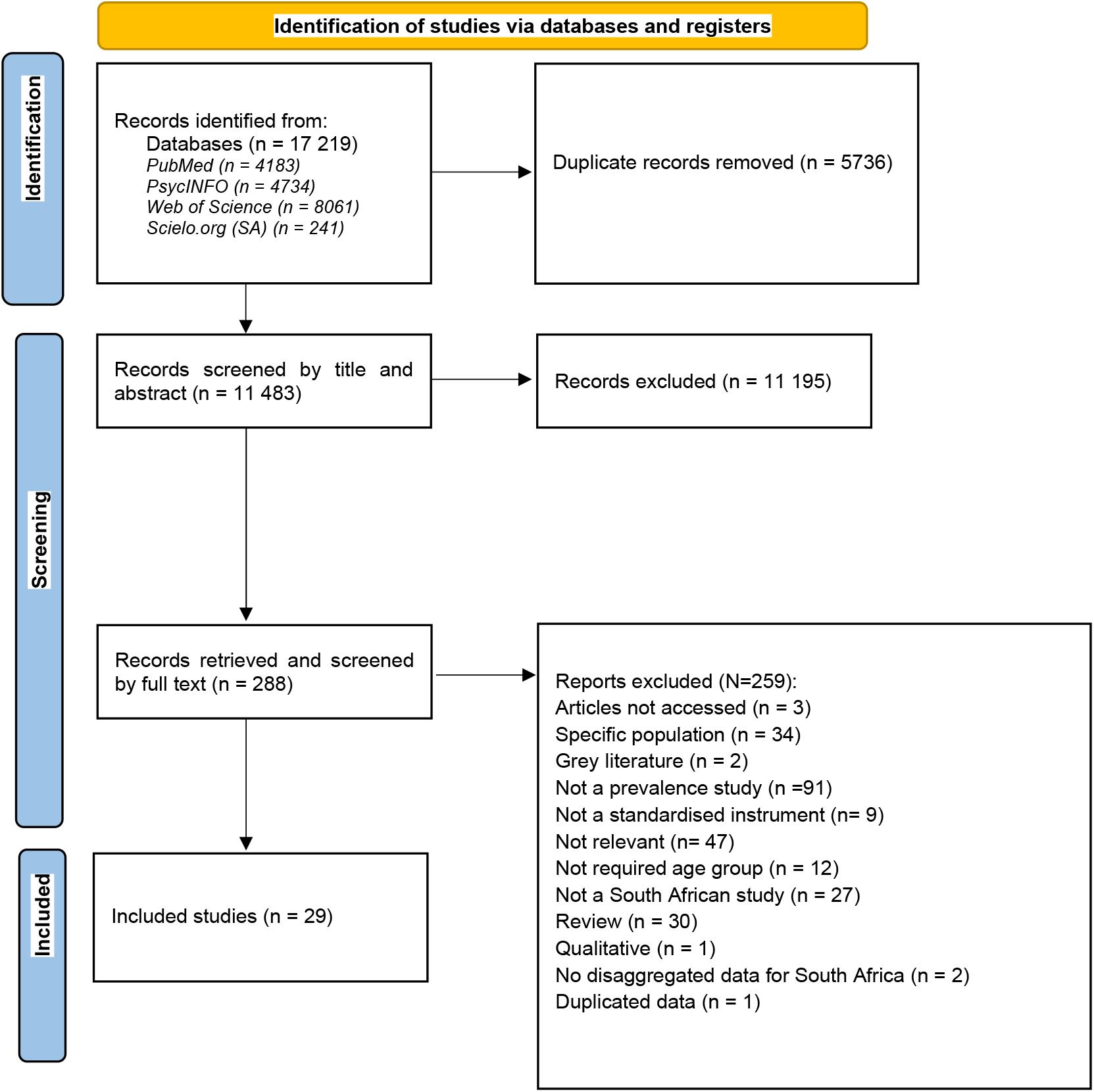
PRISMA flow diagram substance use publications

There was fairly even geographic distribution of studies across South Africa, with 5 national studies and with the most studies reporting data from Limpopo (n=7) Northern Cape (n=6) and the remaining provinces having 5 studies each (see Figure 2) (some studies reported data from more than one province).

**Figure 2:**
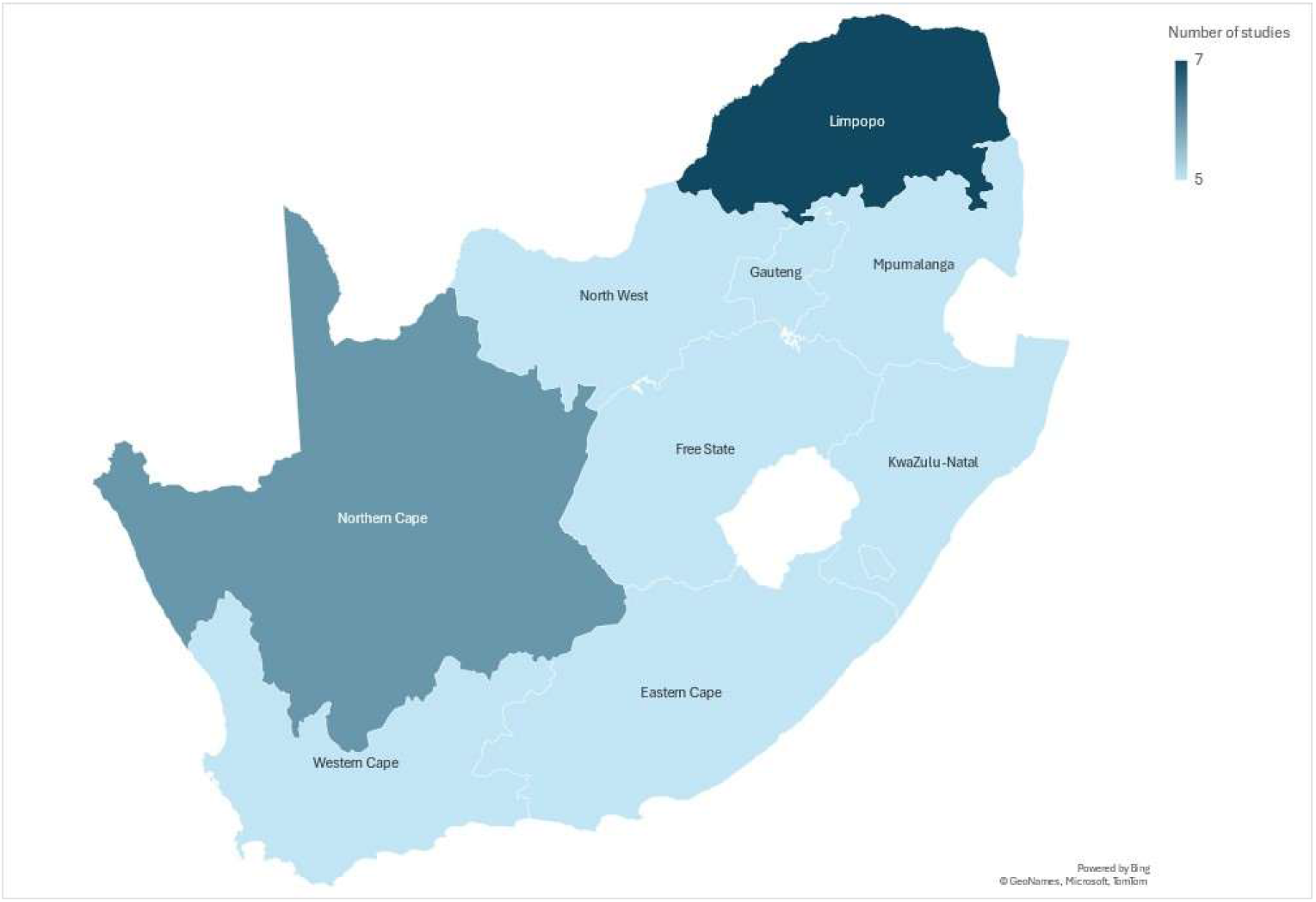
Heat map showing geographic distribution of studies across the 9 provinces in South Africa (n=29).

There were 202 unique prevalence estimates for substance use, across 30 studies. The most investigated substances (higher number of studies, K) based on lifetime and 12-month use as defined in our methods, were tobacco (K=36, N=101,796), alcohol (K=37, N=89,187), and cannabis (K=30, N=78,686) and the least commonly investigated substances for lifetime use were amphetamines (5 studies), over the counter drugs (3 studies), tranquilisers (6 studies), Tik (7 studies), club drugs/MDMA (3 studies), and mandrax (4 studies). Nyaope, also known as whoonga, was covered in two studies and is a highly addictive and street drug commonly found in South Africa which is now known to be made up of low grade heroin [33], and these prevalence estimates were included in the meta-analysis of heroin use. Other substance use outcomes reported in only one study were psychedelic use, any substance use, and illicit drug use. All these outcomes reported in less than five studies were excluded from further analysis.

Prevalence estimates were variously reported for life-time substance use, as well as use in the past 12-months, 6-months, 3-months, 1-month and 1-week, making it difficult to assimilate data meaningfully. Our pragmatic approach was to group prevalence estimates into two categories based on the time frame of assessment: lifetime prevalence and 12-month prevalence. The 12-month category also included studies reporting prevalence over shorter periods, such as 6-month and 3-month timepoints, to allow for consistent synthesis across comparable recall periods. Examination of the data also revealed a number of studies that reported binge/hazardous drinking in the 1-month period, so a separate meta-analysis was conducted with these data.

The results of the meta-analysis for lifetime substance use are presented in Table 1. Lifetime prevalence of any substance was 17.11% (95%CI= 13.51-20.72, K=84, n=259 497). The substances with the most frequent lifetime use were alcohol (39.37%, 95%CI=30.36-48.39, K=16, n=11 620), tobacco (25.66% 95%CI=17.12-34.19, K=14, n=8 721), and cannabis (12.63% 95%CI=7.38-17.88, K=10, n=5 793). However, there was significant heterogeneity across included studies for alcohol (I^2^=99.39%, Q(15)= 3418.2713, p<0.0001), tobacco (I^2^=99.04%, Q(13)= 1568.7937, p< 0.0001), and cannabis (I^2^=97.74%, Q(9)= 415.0807, p<0.0001). As seen in Table 1, heterogeneity was high for all substances with 5 or more studies, limiting the reliability of the pooled lifetime prevalence estimates.

The results of the meta-analysis for studies reporting substance use in the past 12-months or less are presented in Table 2. Prevalence of any substance use was 14.25% (95%CI=8.64-19.86, K=34, n=2 333). For substances with 5 or more studies presented in meta-analysis, the most common substances used in the past 12-months were alcohol (33.17% 95%CI=19.51-46.82, K=9, n=7 256), tobacco (15.82% 95%CI=8.73-22.90, K=6, n=5058) and cannabis (8.27% 95%CI=3.96-12.57, K=5, n=4 290). There was, however, also significant heterogeneity across these studies for alcohol (I^2^=99.70%, Q(8)= 2717.9350, p<.0001), tobacco (I^2^=98.10%, Q(5)= 386.3023, p<0.0001) and cannabis (I^2^=96.82%, Q(4)= 196.5977, p< 0.0001).

1-month binge drinking prevalence was 18.11% (95%CI=9.85-26.37, K=5, n=32 546) again with high levels of heterogeneity (I^2^=99.68%, Q(4)= 979.9347, p<.0001). It is noteworthy that there was significant heterogeneity across all substances that had 5 or more studies/prevalence estimates and for which meta-analysis was conducted. Forest plots for lifetime, and 12 month substance use, and S3 and 1 month hazardous drinking are in Figure 3.

**Figure 3.**
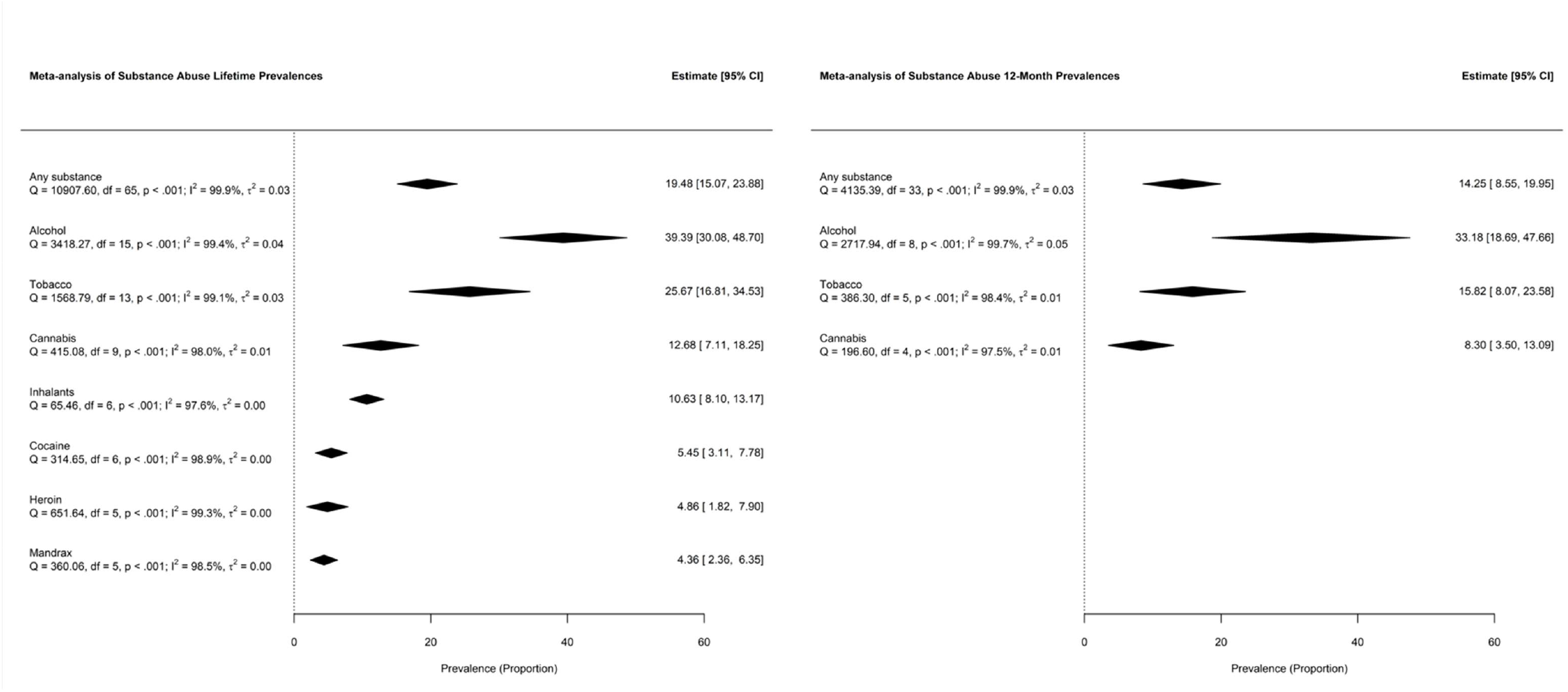
Meta-analysis of lifetime and 12 month substance use prevalence

## Discussion

This review has presented the first synthesized evidence of pooled prevalence of use of the variety of substances used by adolescents in South Africa with our data showing use at concerning levels aligning with the contribution of substance use in the global burden of disease [34]. The findings highlight troubling patterns of exposure to a range of substances including alcohol, tobacco, cannabis, amphetamines, and inhalants. Both lifetime 12-month prevalence data from this review suggest that early initiation of use and exposure to alcohol, tobacco and cannabis are public health concerns for South Africa. Regarding substances most used, alcohol, tobacco, and cannabis showed consistently high lifetime and past-year prevalences. One-month hazardous drinking prevalence of 18% and lifetime alcohol use of 39% are alarmingly high exposures and raise the public health concern of the pathway to development of substance use disorders later in life. Exposure to environmental stressors, poverty and violence, and overall low wellbeing, which are an ongoing reality for many young people in South Africa is closely related to use of alcohol and tobacco by adolescents in SA [18] and data from this review underscore the potential influence of psychosocial determinants in substance use patterns urgent need to address substance use in young people in SA.

Previous evidence has indicated early initiation of alcohol use and increasing binge drinking among young people in SA particularly young women [35]. These levels of exposure may be linked to exposure to alcohol marketing among adolescents in SA [36]. Tobacco prevalence from this review is similarly high, and aligns with findings from a previous SA review on adolescent tobacco use which found a 22% prevalence of use among adolescents and young adults [37], and importantly this does not yet capture the exposure to new tobacco products, including e-cigarettes. E-cigarette consumption is an emerging concern, with 17.14% lifetime use estimated in 2019.in Brazil (Marmirolli FAP, Garcia VMPS, Fidalgo TM. E-cigarette use in a nationally representative sample of adolescents. Int JMent Health Addict 2024; published online July 12. DOI:10.1007/s11469-024-01363-4). Lifetime exposure to inhalants in this study was >10% suggesting experimentation extends beyond socially normalized substances.

The prevalence for one month alcohol use estimate from this review (in comparison with SA 33.17% 12-month use) Data from recent Global School-based Student Health Survey One month alcohol use 25.2% {Farnia, 2024 #88} and nationally representative data from Brazil (21.2%) different estimates 29.3% {PSICOTRÓPICAS, #89} lifetime 59.6 {Bloch, 2015 #90} Although direct comparisons across countries are not possible with this data, these prevalence rates for SA exceed those reported in other Sub-Saharan African (SSA) countries. Regional estimates across eight SSA countries showed past-month cannabis use (4.39%) and lifetime amphetamine use (3.05%), lower than SA levels. Nationally representative data from Ghana on alcohol use behaviours indicated 12.6% current alcohol use [38] (in comparison with SA 33.17% 12-month use). Interestingly, SA’s geographical neighbour (with similar sociocultural context) has similar use data for alcohol, from nationally representative data. This shows 29.8% past-month alcohol use and prevalence of lifetime drunkenness of 26.0% [39].

The heterogeneity of the included studies presents an important limitation to the findings of this review. Variability in study designs, population characteristics, sample sizes, outcome measures, and data collection methods contributes to inconsistencies in results, making direct comparisons challenging. As a result, the generalizability of these findings is limited, the pooled estimates are interpreted with caution, and the body of evidence points to the need for more robust estimates. Future research should aim for greater standardization in measurement, definitions of substance use categories and inclusion of underrepresented substances, such as methamphetamine (tik), MDMA, and prescription medications which all have potential for harm and dependence.

Previous SA studies have underscored the gaps that remain in data for understanding the burden of substance use in SA, particularly for newer psychoactive substances and rural populations. For more than a decade there have been calls for more robust and nationally representative prevalence estimates to inform targeted interventions [40]. With this review, we reinforces this pressing need and call for key areas of future research and surveillance. Robust and nationally representative evidence on prevalence of substance use is needed to enable translational epidemiology [41] for scaled up implementation of prevention and treatment programmes. Standardized surveillance systems, longitudinal studies, and the integration of biological and self-reported data are needed to enhance the accuracy of prevalence estimates and improve the design of tailored interventions for young people at risk. Reliable epidemiological data help identify patterns and trends in substance use, assess the burden on public health systems, and allocate resources efficiently [42]. Without comprehensive and up-to-date prevalence estimates, interventions may be misdirected, failing to target the most vulnerable populations or emerging substances of concern [15] and failing to make most effective use of resources for health, which SA can not afford to do. Longitudinal research similarly will enable tracking of substance use patterns and risk factors over time. Brazilian review also highlights an approach for a nationally representative survey with over 200.000 adolescents, which we could point as a potential methodology in the need of strenghtening SA data {Marchionatti, 2024 #53} This evidence is critical for evaluating the long-term effectiveness of interventions, identifying emerging trends, and tailoring prevention and treatment strategies to the evolving needs of the population.

## Conclusion

This review presents the first comprehensive synthesis of substance use among South African youth, highlighting concerningly high prevalence of alcohol, tobacco, and cannabis use as well as use of other substances with less data. The review has limitations stemming from the variation in the included studies, limiting the useful synthesis of the data available. Consequently the findings underscore the urgent need for nationally representative survey data to accurately present the scale of this important public health need, and guide policy and programmes for reducing the harms of substance use in young people. Crucially, evidence-based prevention strategies, particularly those targeting adolescents during the early stages of substance use, will be informed by this representative data and are critical to preventing progression to substance use disorders.

## Data Availability

All data produced in the present work are contained in the manuscript

